# SARS-CoV-2 antibodies, serum inflammatory biomarkers and clinical severity of hospitalized COVID-19 Patients

**DOI:** 10.1101/2020.07.22.20159673

**Authors:** Roberto Gozalbo-Rovira, Estela Gimenez, Víctor Latorre, Clara Francés-Gómez, Eliseo Albert, Javier Buesa, Alberto Marina, María Luisa Blasco, Jaime Signes-Costa, Jesús Rodríguez-Díaz, Ron Geller, David Navarro

**Author notes:** **Corresponding authors** David Navarro, Microbiology Service, Hospital Clínico Universitario, Instituto de Investigación INCLIVA, Valencia, and Department of Microbiology, University of Valencia, Valencia, Spain. Av. Blasco Ibáñez 17, 46010 Valencia, Spain., Ron Geller: Institute for Integrative Systems Biology (I2SysBio), Universitat de Valencia-CSIC, 46980 Valencia, Spain, Jesús Rodríguez-Díaz: Department of Microbiology, Faculty of Medicine, University of Valencia, AV. Blasco Ibañez 17, 46010, Valencia. Spain. **Equal contributors** (Roberto Gozalbo-Rovira, Estela Gimenez, Víctor Latorre, Clara Francés-Gómez). **Article’s main point:** The levels of neutralizing antibodies (NtAb) against the SARS-CoV-2 spike protein and IgGs targeting its receptor binding domain were comparable at different time points after the onset of COVID-19 between patients admitted to ICU or the pneumology ward. Weak or very weak correlations were found between serum levels of these antibody responses and those of several biomarkers such as CRP, ferritin, LDH, Dimer-D, or IL-6, known to behave as surrogates for COVID-19 severity.

## Abstract

**Background:** The involvement of SARS-CoV-2 antibodies in mediating immunopathogenetic events in COVID-19 patients has been suggested. By using several experimental approaches, we investigated the potential association between SARS-CoV-2 IgGs recognizing the spike (S) protein receptor-binding domain (RBD), neutralizing antibodies (NtAb) targeting S, and COVID-19 severity.

**Patients and Methods:** This unicenter, retrospective, observational study included 51 hospitalized patients (24 at the intensive care unit; ICU). A total of 93 sera from these patients collected at different time points from the onset of symptoms were analyzed. SARS-CoV-2 RBD IgGs were quantitated by ELISA and NtAb_50_ titers were measured in a GFP reporter-based pseudotyped virus platform. Demographic and clinical data, complete blood counts, as well as serum levels of ferritin, Dimer-D, C reactive protein (CRP), lactose dehydrogenase (LDH), and interleukin-6 (IL-6) were retrieved from clinical charts.

**Results:** The overall correlation between levels of both antibody measurements was good (Rho=0.79; *P*=0<0.001). SARS-CoV-2 RBD IgG and NtAb_50_ levels in sera collected up to day 30 after the onset of symptoms were comparable between ICU and non-ICU patients (*P*=>0.1). The percentage of patients who exhibited high NtAb_50_ titers (≥ 160) was similar (*P*=0.20) in ICU (79%) and non-ICU (60%) patients. Four ICU patients died; two of these achieved NtAb_50_ titers ≥ 1/160 while the other two exhibited a 1/80 titer. Very weak (Rho=>0.0-<0.2) or weak (Rho=>0.2-<0.4) correlations were observed between anti-RBD IgGs, NtAb_50,_ and serum levels pro-inflammatory biomarkers.

**Conclusions:** The data presented herein do not support an association between SARS-CoV-2 RBD IgG or NtAb_50_ levels and COVID-19 severity.

## INTRODUCTION

Coronavirus disease 2019 (COVID-19), caused by severe acute respiratory syndrome coronavirus 2 (SARS-CoV-2), emerged in late 2019 and has been declared a pandemic [1]. Clinical presentation of COVID-19 varies widely, ranging from asymptomatic to mild or severe forms [2,3]. Worse clinical outcomes are related to an imbalanced immune response skewed toward a Th_1_ pro-inflammatory profile, which leads to the uncontrolled release of cytokines and chemokines, such as interleukin-6 (IL-6), that mediates progression into acute respiratory distress syndrome, multiorgan failure, and death [4,5].

Adaptive humoral immunity is thought to protect from acquiring SARS-CoV-2 infection, of which neutralizing antibodies (NtAb) seemingly play a major role [6]. Although epitopes mapping within all SARS-CoV-2 structural proteins have been shown to elicit NtAb, the receptor-binding domain (RBD) of the viral spike protein (S) is immunodominant and a highly specific target of most potent NtAbs in COVID-19 patients [6-9]. The involvement of functional antibodies in SARS-CoV-2 clearance and modulation of COVID-19 severity remains to be precisely defined [10]. Data obtained in experimental models indicated that adoptive transfer of neutralizing monoclonal antibodies reduces viral burden in the lung, ameliorates local inflammation and decreases mortality [7,11,12]. Moreover, passive immunization of critically ill COVID-19 patients with plasma from individuals who had recovered from SARS-CoV-2 infection and seroconverted was associated with improved clinical outcomes in uncontrolled case series [13,14]. Yet, the possibility that antibodies could potentially trigger immunopathogenic events in SARS-CoV-2-infected patients or enhance infection is a major concern [6,15,16]. In this context, higher antibody titers, either neutralizing or not, have been reported to be present in patients developing severe forms of COVID-19 when compared to mildly symptomatic individuals who did not require hospitalization [17-23]. Here, we aimed to explore the potential relationship between the magnitude of SARS-CoV-2 antibodies binding to RBD and NtAb targeting the S protein with the severity of COVID-19 in a cohort of hospitalized patients.

## PATIENTS AND METHODS

### COVID-19 patients

In this unicenter, retrospective observational study, 51 non-consecutive patients with laboratory-confirmed SARS-CoV-2 infection by RT-PCR, admitted to Hospital Clínico Universitario of Valencia between March 5 to April 30, 2020, were included. The availability of leftover cryopreserved sera for the experiments detailed below was the only inclusion criterium. Out of the 51 patients in this series, 27 were hospitalized in the pneumology ward and 24 in the intensive care unit (ICU), of whom 16 underwent mechanical ventilation and 4 eventually died. Patients were hospitalized within 24 h after seeking medical attention at the emergency service. All patients presented with pneumonia and imaging/laboratory findings compatible with COVID-19 [2,3]. Patients admitted to ICU had severe respiratory compromise, defined by failure to maintain an arterial oxygen saturation of >90% despite receiving supplemental oxygen at 50%, and/or a respiratory rate greater than 35 breaths per minute. Medical history and laboratory data were retrospectively reviewed. The study period for each patient comprised the time from hospitalization to discharge or death. The current study was approved by the Research Ethics Committee of Hospital Clínico Universitario INCLIVA (March, 2020).

### Patient Samples

A total of 93 sera from 51 patients with COVID-19 were included for the analyses detailed below. Forty-seven sera were obtained within the first two weeks after the onset of symptoms, 32 between the third and the forth weeks and 14 afterwards (between days 31 and 45). Sequential specimens were available from 20 out of the 51 patients (median 3 specimens/patients; range 2 to 6), 17 of whom were in ICU. Sera from 51 individuals collected prior to the epidemic outbreak (within years 2018 and 2019) served as controls in the SARS-CoV-2 RBD IgG immunoassay and the SARS-CoV-2 neutralizing antibody assays described below. Nine patients had tested positive for Coronavirus 229E by the xTAG Respiratory Viral Panel (Luminex Corporation, Austin, Tx, USA).

### SARS-CoV2-2 RT-PCR

Nasopharyngeal or oropharyngeal specimens were obtained with flocked swabs in universal transport medium (Beckton Dickinson, Sparks, MD, USA, or Copan Diagnostics, Murrieta, CA, USA) and conserved at 4 °C until processed (within 6 hours). Undiluted tracheal aspirate samples obtained from mechanically ventilated patients were also processed when available. Commercially-available RT-PCR kits were used for SARS-CoV-2 RNA testing, as previously detailed [24].

### SARS-CoV-2 RBD IgG immunoassay

An enzyme-linked immunosorbent assay (ELISA) was used to quantitate IgG antibodies binding to SARS-CoV-2 RBD [25]. A detailed description of the assay can be found in Supplementary Methods. Briefly, SARS-CoV-2 RBD was produced in Sf9 insect cells infected with recombinant baculoviruses (Invitrogen, CA, USA). Following purification, the protein was concentrated to 5 mg/mL by ultrafiltration. Ninety-six well microplates were coated with RBD at 1 μg/mL. Serum samples were diluted 1:500 in phosphate-buffered saline-Tween (PBS-T) containing 1% bovine serum albumin and run in triplicate (mean values are reported). The plates were incubated with 1:5,000 dilution of horseradish peroxidase (HRP)-conjugated goat anti-human IgG (Jackson Laboratories). After three washes with PBS-T, the binding was detected using SigmaFast OPD reagent (Sigma) according to manufacturer’s recommendation. Color development was stopped with 3M H_2_SO_4_ and read on a Multiskan FC (ThermoFischer Scientific) plate reader at 492 nm. Serial sera from individual patients were analyzed in the same run. The cut-off discriminating between positive and negative sera was set as the mean absorbance of control sera plus three times the standard deviation. SARS-CoV-2 RBD IgG avidity index was calculated as the percentage of measured optical density (OD) in 6M urea-treated wells relative to that in the untreated wells: AI (%) = OD of urea-treated well × 100/OD of non-urea-treated well [26]. A positive-control (high avidity) specimen derived from a convalescent-phase serum from a COVID-19 patient (AI, 84%) was included on each ELISA plate.

### SARS-CoV-2 neutralizing antibody assay

A green fluorescent protein (GFP) reporter-based neutralization assay which used a non-replicative vesicular stomatitis virus pseudotyped with the SARS-CoV-2 spike protein (VSV-S) was optimized as previously described (see supplementary methods) [27-29]. Neutralization assays were performed on Vero cells. Sera were heat-inactivated for 30 minutes at 56°C then brought to an initial dilution of 1/10, followed by four 4-fold dilutions in duplicate. Each dilution was mixed with an equal volume containing 1,250 focus forming units of the VSV-S virus and incubated at 37°C for 1 h. The mixture was then added to Vero cells in 96-well plates and incubated for 18 hours, after which GFP expression was measured using a live cell microscope system (IncuCyteS3, Sartorious). Background fluorescence from uninfected cells was subtracted from all values, followed by standardization to the average GFP expression of mock-treated, infected cells. All sera which did not reduce viral replication by 50% at a 1/20 dilution were considered non-neutralizing and were arbitrarily assigned a value of 1/10. All sera that did not result in >70% recovery of GFP signal at the highest antibody dilution were retested using 5-fold dilutions ranging between 100 and 12,500-fold. Finally, the lowest antibody dilution resulting in >50% virus neutralization was used as the NtAb_50_ value. Here, we considered high NtAb_50_ titers those ≥ 1/160, as this is the minimum NtAb titer of plasma from COVID-19 convalescent individuals recommended by the FDA for therapeutic use [30].

### Laboratory measurements

Clinical laboratory investigation included complete blood count and levels of ferritin, Dimer-D, C reactive protein (CRP), lactose dehydrogenase (LDH) and interleukin-6 (IL-6) quantitated in sera that were later used for SARS-CoV-2 RBD IgGs and NtAb testing.

### Statistical methods

Frequency comparisons for categorical variables were carried out using the Fisher exact test. Differences between medians were compared using the Mann–Whitney U-test. Spearman’s rank test was used to assess the correlation between continuous variables using the entire dataset (i.e. individuals with single and repeated measurements). Receiver operating characteristic (ROC) curve analysis was performed to identify the optimal SARS-CoV-2 RBD IgG level predicting NtAb titers above a certain threshold. Two-sided exact *P*-values are reported. A *P*-value <0.05 was considered statistically significant. The analyses were performed using SPSS version 20.0 (SPSS, Chicago, IL, USA).

## RESULTS

### Clinical characteristics of COVID-19 patients

Patients hospitalized in the pneumology ward (n=27) and ICU (n=24) were matched for sex and age, the presence of co-morbidities and the time elapsed from the day of onset of symptoms to first serum sample collection (Table 1). As expected, ICU patients were hospitalized for longer periods. Median serum levels of several pro-inflammatory biomarkers, such as LDH, dimer-D and IL-6, were significantly higher in ICU patients than in non-ICU patients, further confirming their association with COVID-19 severity [2-5]. In contrast, the median total lymphocyte counts did not differ across comparison groups (Table 1).

**TABLE 1.** Demographic, clinical and laboratory characteristics of patients with COVID-19.

|  |  |  |  |  |
| --- | --- | --- | --- | --- |
| **Parameter** | **All patients** | **Patients hospitalized in the pneumology ward** | **Patients hospitalized in the intensive care unit** | **P value** |
| **Sex: Male/Female; no. (%)** | 32 (63)/<br>19 (37) | 14 (52)/ 13 (48) | 18 (75)/ (6 (25) | 0.15 |
| **Age; median (range)** | 53 (21-77) | 58 (42-76) | 65 (29-77) | 0.07 |
| **Days of hospitalization; median (range)** | 17 (2-67) | 9 (2-22) | 36 (8-67) | <0.001 |
| **Days from onset symptoms to first serum sample; median (range)** | 12 (5-36) | 11 (5-32) | 13 (7-36) | 0.33 |
| **Co-morbidities; no. (%)** | 35 (69) | 18 (67) | 17 (71) | 0.75 |
| **Number of comorbidities; median (range)** | 1 (0-5) | 1 (0-3) | 2 (0-5) | 0.18 |
| **Comorbidity; median (range)** |  |  |  |  |
| Arterial hypertension | 23 (45) | 11 (41) | 12 (50) | 0.58 |
| Chronic renal disease | 2 (4) | 0 | 2 (8) | 0.22 |
| Diabetes mellitus | 12 (24) | 5 (19) | 7 (29) | 0.51 |
| Dyslipidemia | 16 (31) | 7 (26) | 9 (38) | 0.37 |
| Ischemic cardiovascular disease | 4 (8) | 2 (7) | 2 (8) | 0.90 |
| Myocardial infarction | 2 (4) | 1 (4) | 1 (4) | 1.00 |
| Pulmonar disease<sup>a</sup> | 7 (14) | 2 (7) | 5 (21) | 0.16 |
| Tumor | 3 (6) | 1 (4) | 2 (8) | 0.48 |
| **Laboratory findings<sup>b</sup>; median (range)** |  |  |  |  |
| CRP (in mg/l) | 44 (0.8-273) | 70 (0.8-242) | 24.80 (1.00-273) | 0.24 |
| Ferritin (ng/ml) | 674 (2.5-2986) | 565 (9.2-2779) | 959 (2.50-2986) | 0.17 |
| Dimer-D (ng/ml) | 903 (91-5445) | 488 (91-1894) | 1328 (489-5445) | <0.001 |
| LDH (U/l) | 666 (357-1328) | 556 (357-825) | 790 (518-1328) | <0.001 |
| IL-6 (pg/ml)<sup>c</sup> | 1012 (4.6-5000) | 79 (4.6-124) | 1277 (186-5000) | 0.009 |
| Total lymphocyte count (*10<sup>9</sup>/L) | 1.15 (0.17-3.98) | 1.13 (0.17-2.95) | 1.31 (0.38-3.98) | 0.17 |
| <sup>a</sup>Including asthma, atelectasis and chronic obstructive pulmonary disease.<br><sup>b</sup>The median was calculated in patients with more than one sample.<br>Normal values: 12-300 ng/ml for ferritin, <100 ng/ml for Dimer-D, and <10 mg/L for C-reactive protein (CRP), 140-280 U/L Lactic acid dehydrogenase (LDH), 5-15pg/ml for IL-6, and 1-4.8 lymphocytes x10<sup>9</sup>/L.<br><sup>c</sup>Data available from 18 patients. |  |  |  |  |

### Correlation between SARS-CoV-2 RBD IgG levels and neutralizing antibody titers

We first aimed to determine whether SARS-CoV-2 RBD IgGs quantified by ELISA could be used as a proxy for NtAb_50_ titers, as measured in a reporter-based SARS-CoV-2 spike protein pseudotyped VSV neutralization platform. As shown in Figure 1, the overall correlation between levels of both antibody assays was fairly good (Rho=0.79; *P*<0.001). ROC analysis showed that SARS-CoV-2-RBD IgG levels ≥ 2.34 AU/ml predicted the presence of NtAb_50_ titers ≥ 160 with a sensitivity of 84% and a specificity of 95% (Supplementary Figure 1).

**Figure 1.**
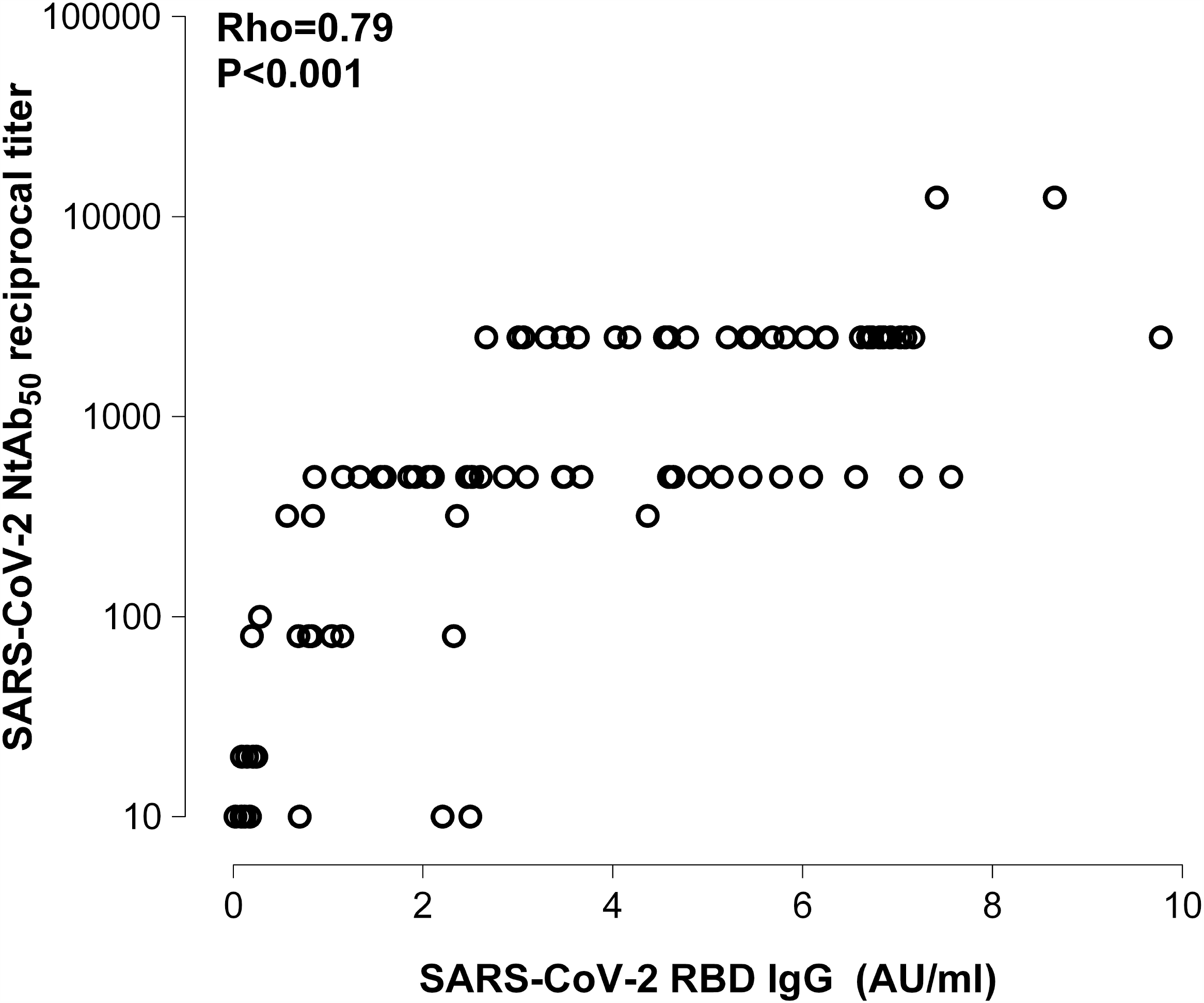
Correlation between SARS-CoV-2 RBD IgG levels quantitated by ELISA and NtAb_50_ titers measured by a reporter-based pseudotype (VSV-S) neutralization assay in sera from COVID-19 patients. Rho and P values are shown.

### Kinetics of SARS-CoV-2 RBD IgGs and neutralizing antibodies

SARS-CoV-2 RBD IgGs and NtAb_50_ levels at different times after the onset of symptoms are shown in Figure 2. Overall, serum levels of both antibody tests were seen to increase significantly in parallel over time, although the median peak NtAb_50_ titer was reached earlier (between days 11-20) than that of RBD-specific IgGs (between days 20-30). After peaking, NtAb_50_ levels remained stable through the end of the study period, while RBD-specific IgGs decreased slightly afterwards. Sequential sera were available from 20 patients, most of whom (n=17) were at ICU. The kinetics profile from both antibody assays was found to vary widely across patients (Figure 3), some of whom exhibited increasing levels while others displayed either constant or fluctuating titers.

**Figure 2.**
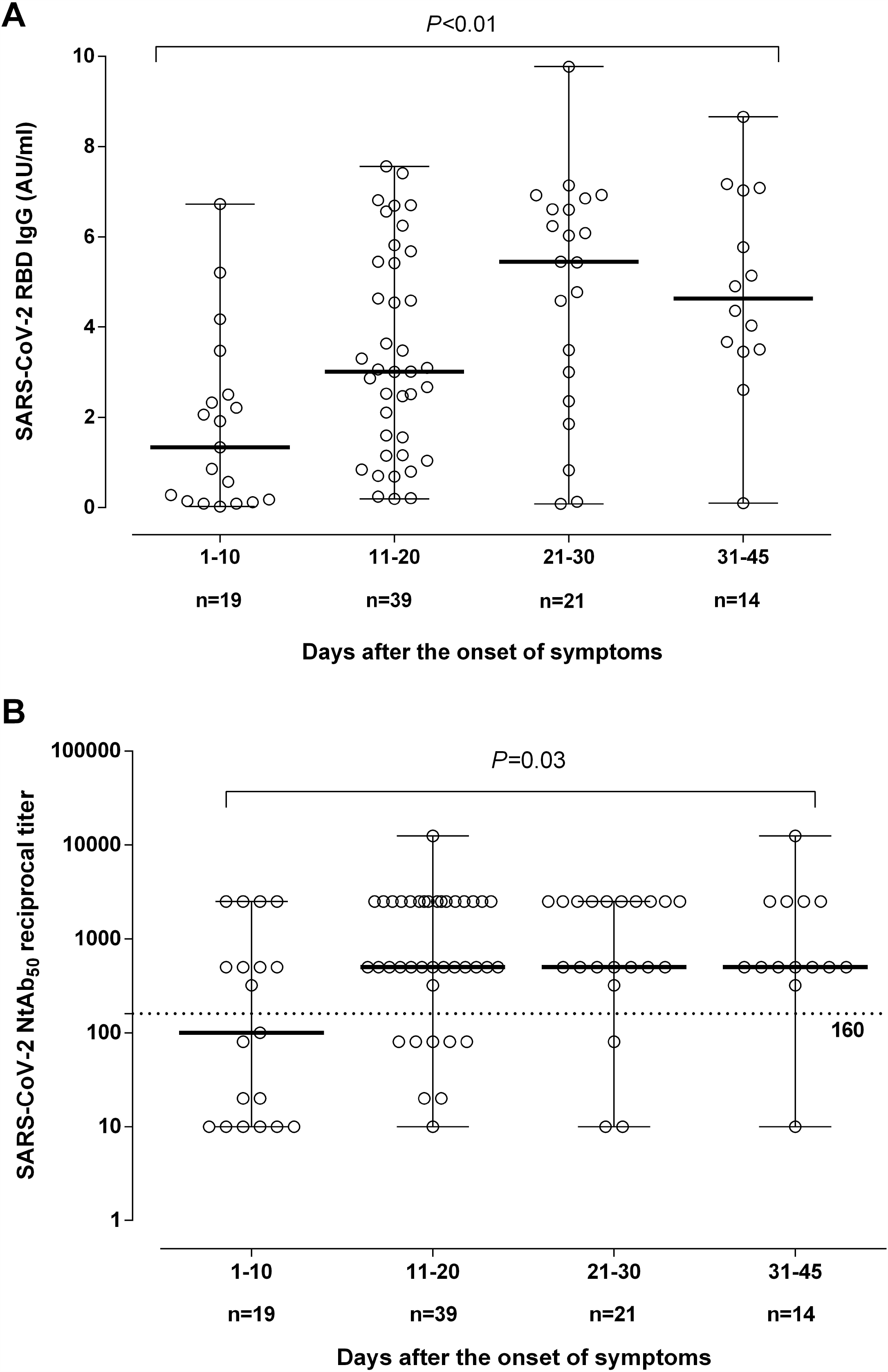
SARS-CoV-2 RBD IgG levels (A) and NtAb_50_ titers (B) at different time points after the onset of symptoms in patients with COVID-19.

**Figure 3.**
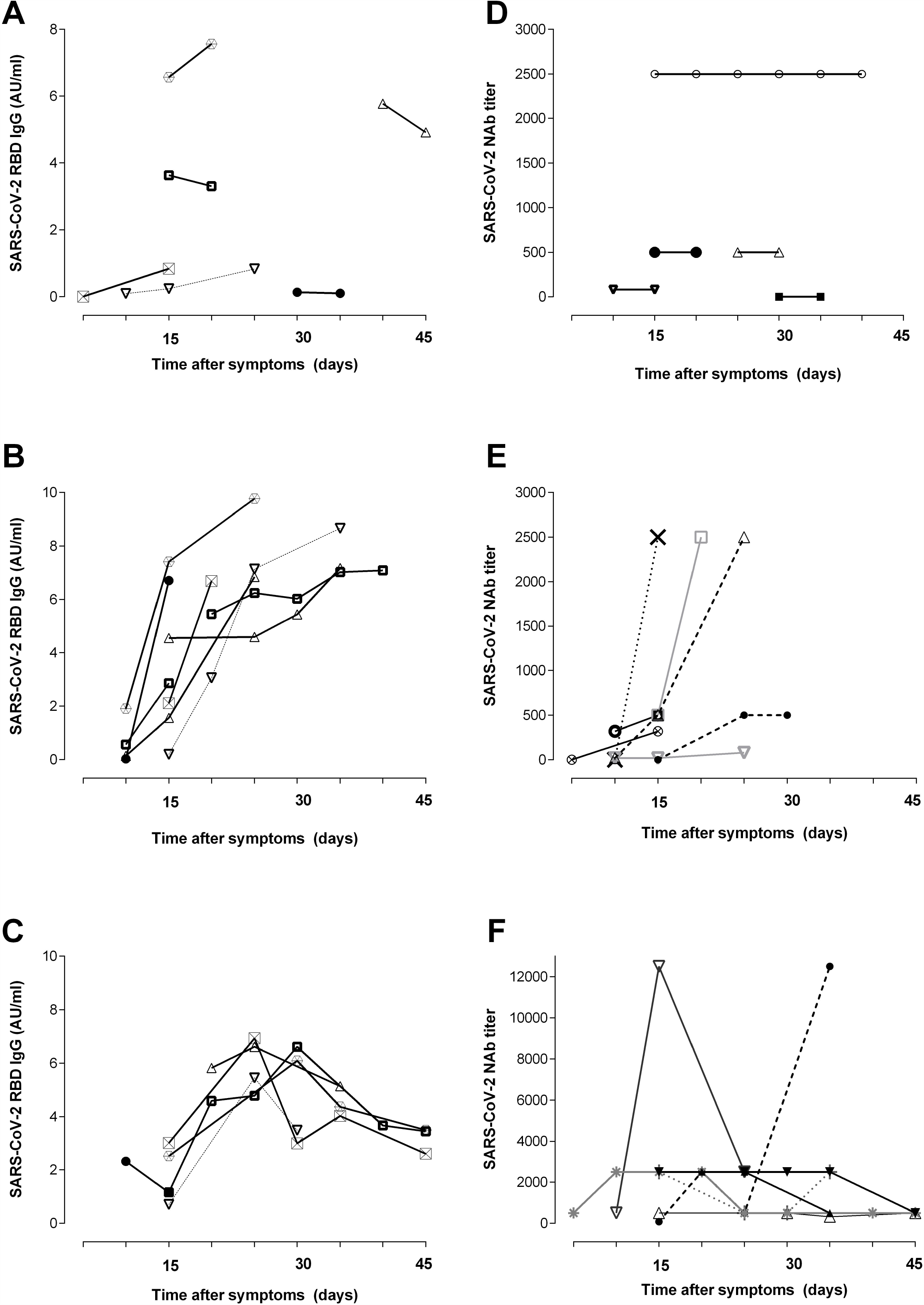
Kinetics patterns of SARS-CoV-2 RBD IgGs (A,B,C) and NtAb (D,E,F) in 20 COVID-19 patients (17 admitted to the intensive care unit).

### SARS-CoV-2 RBD IgG avidity

Avidity of SARS-CoV-2 IgGs in sera from COVID-19 patients was assessed by a conventional urea dissociation assay [26]. Overall, AIs were very low (median 5%; range 2-28%). Most sera (40 out of 51) displayed AI ≤ 10%. Analysis of sequential sera from 20 patients revealed that SARS-CoV-2 IgG AI slightly increased over time (Figure 4). SARS-CoV-2 RBD IgG AI did not correlate with NtAb_50_ titers (Rho=0.07; *P*=0.56)

**Figure 4.**
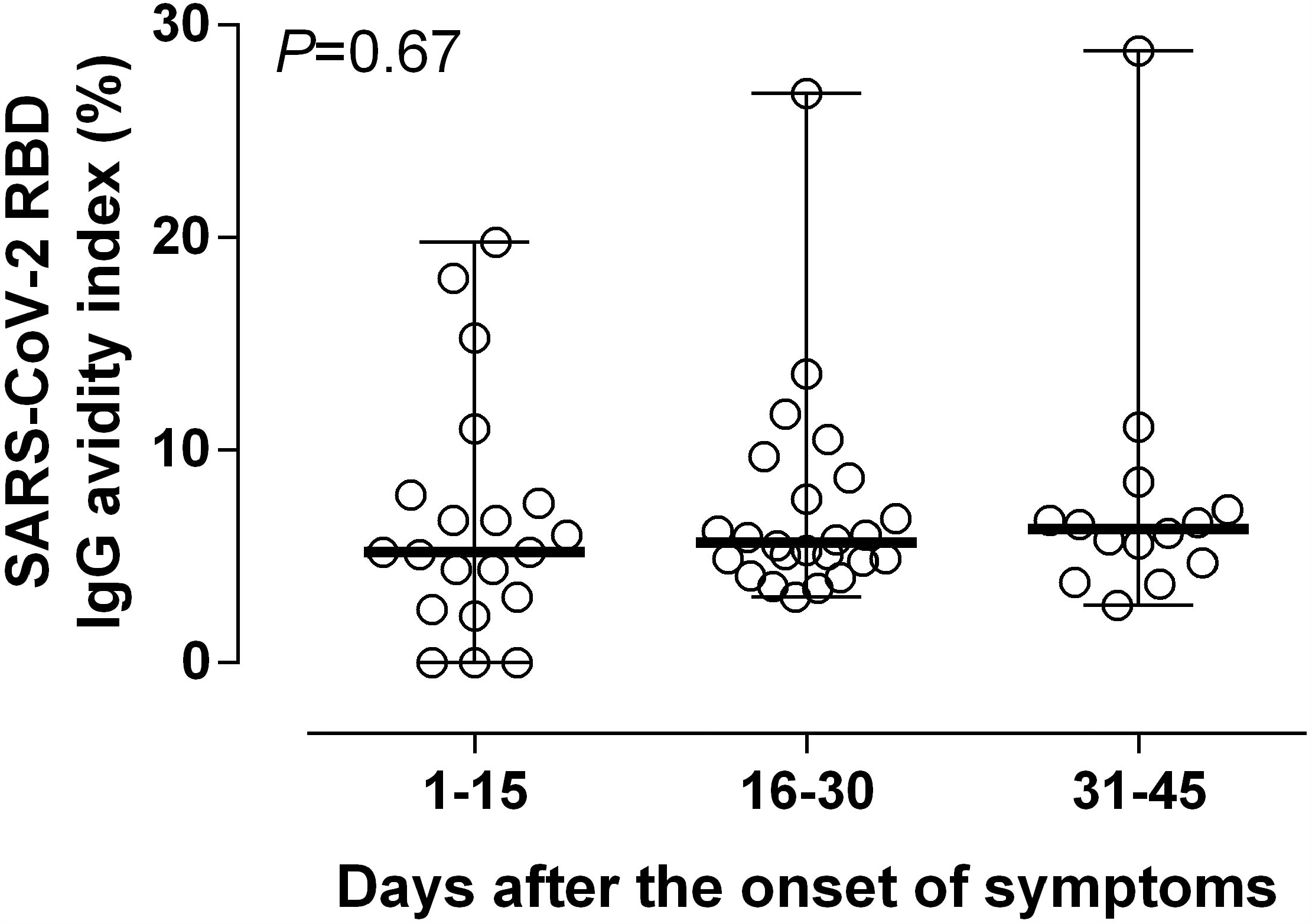
SARS-CoV-2 RBD IgG avidity indices (AIs) of serial sera from COVID-19 patients collected at different times following the onset of symptoms.

### SARS-CoV-2 antibodies and COVID-19 severity

We next compared SARS-CoV-2 RBD IgG and NtAb_50_ levels in ICU and non-ICU patients in sera collected within the first 30 days after the onset of symptoms. We did not notice a significant difference in the magnitude of either antibody response across groups (Figure 5). Comparison between groups at later times was not possible due to the scarce number of sera (n=1) available from non-ICU patients. The percentage of patients who reached NtAb_50_ titers ≥ 160 was comparable (*P*=0.20) in ICU (79%) and non-ICU (60%) patients. Of note, 4 ICU patients died, of which two achieved NtAb_50_ titers ≥ 1/160 while the other two exhibited a 1/80 titer.

**Figure 5.**
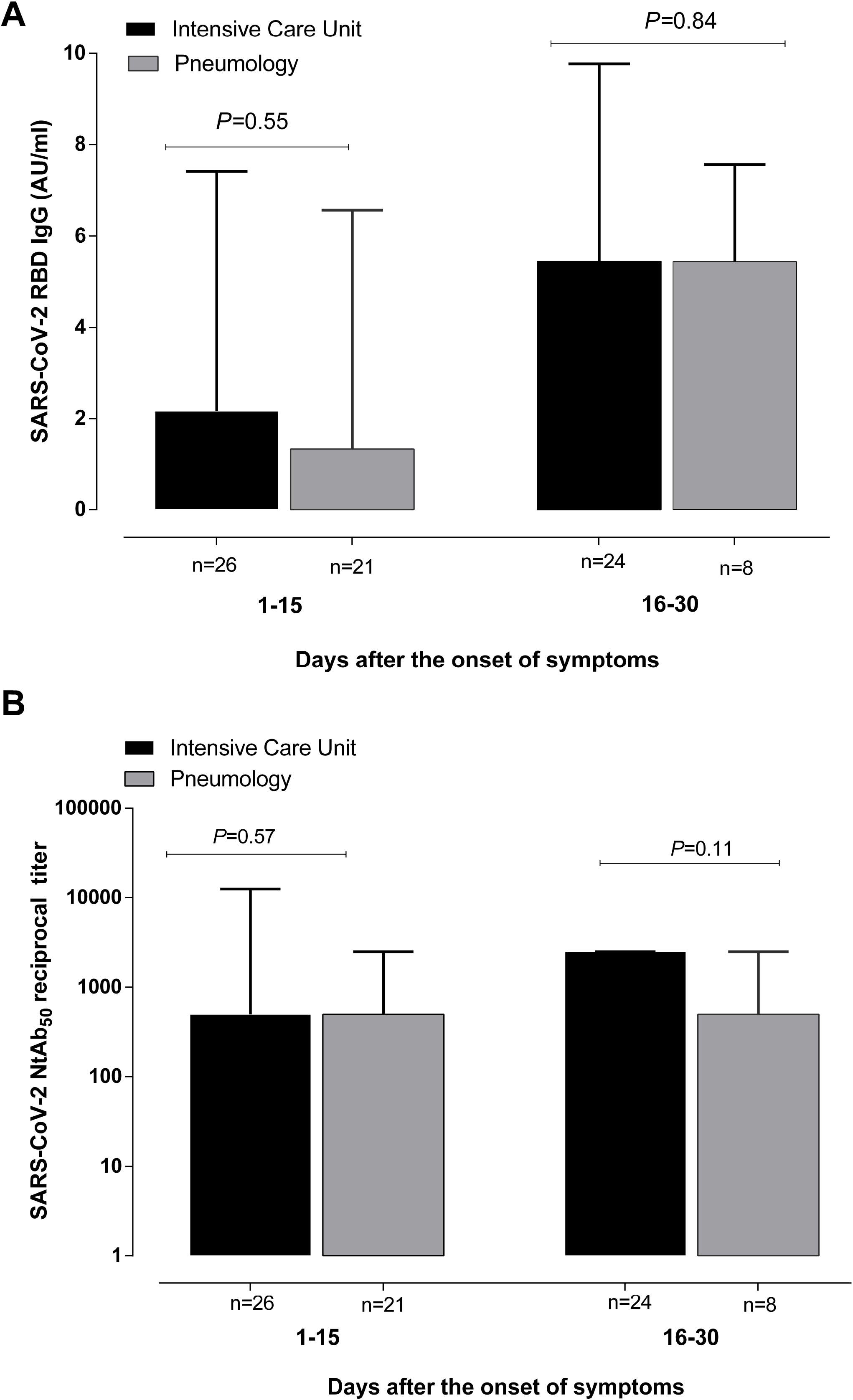
SARS-CoV-2 RBD IgG levels (A) and NtAb_50_ titers (B) at different time points after the onset of symptoms in patients with COVID-19 either admitted to the intensive care unit or the pneumology ward. P values for comparisons are shown.

### SARS-CoV-2 antibody levels and biomarkers of COVID-19 prognosis

Finally, we sought to determine whether the magnitude of SARS-CoV-2 RBD IgG and NtAb responses was related to an inflammatory state, as inferred from serum levels of CRP, ferritin, Dimer-D, LDH and IL-6. For this, we first performed correlation analyses between these parameters. Very weak (Rho=>0.0-<0.2) or weak (Rho=>0.2-<0.4) correlations (either positive or negative) were found between SARS-CoV-2 RBD IgG levels or NtAb_50_ titers and all selected biomarkers when considering the entire data set (Figure 6) or when analyses were done separately for specimens collected at different time frames after the onset of symptoms (days 1-15 or days 15-30; not shown). Measurements from both antibody assays weakly correlated with total lymphocyte counts. As a complementary approach, we grouped sera into two categories (high NtAb_50_ titers: ≥ 1/160 and low NtAb_50_ titers: <1/160), and assessed whether median levels of the abovementioned parameters differed across groups. We found this not to be the case (Supplementary Figure 2).

**Figure 6.**
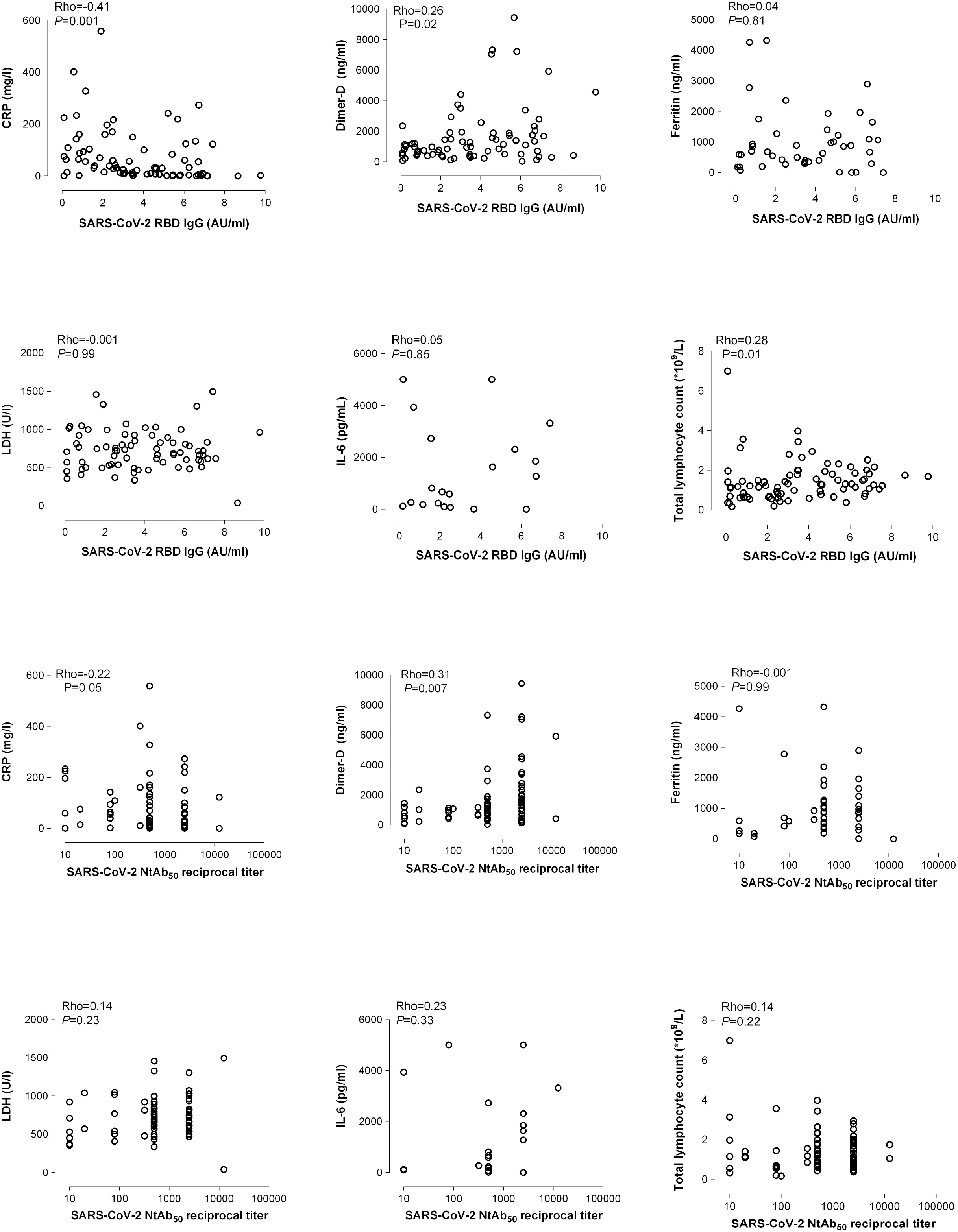
Correlation between SARS-CoV-2 RBD IgG levels and NtAb_50_ titers with serum levels of C-reactive protein (CRP), Dimer-D, ferritin, lactate dehydrogenase (LDH), interleukin-6 (IL-6) and absolute lymphocyte counts. Rho and *P* values are shown.

## DISCUSSION

Here, in addition to further characterizing the antibody response to SARS-CoV-2 in hospitalized COVID-19 patients, we mainly aimed to determine whether a relationship could be established between the magnitude of SARS-CoV-2 RBD IgG and NtAb levels and the “inflammatory state” of patients, which has been shown to directly correlate with COVID-19 severity and prognosis [2-5].

We found that SARS-CoV-2 RBD IgG levels correlated fairly well with NtAb titers, as quantitated by a VSV reporter virus pseudotyped with SARS-CoV-2 S protein (VSV-S), thus lending support to the assumption that the former parameter is a reasonably reliable proxy for the latter. This was expected as RBD encompasses the most critical region of SARS-CoV-2 for ACE2 receptor binding [8,9]. Moreover, we could define a SARS-CoV-2 RBD IgG threshold (≥ 2.34 AU/ml) predicting NtAb titers ≥ 1/160 with high sensitivity and specificity, this being the lowest titer of plasma recommended by FDA for passive transfer therapy [30].

Previous studies have reported a correlation between RBD IgG levels and NtAb titers in patients with comparable or less severe clinical presentations of COVID-19, using either live native SARS-CoV-2 virus, engineered SARS-CoV-2 pseudotype virus systems or replication-competent SARS-CoV-2 chimeric viruses [18,22,30-36]. The degree of correlation between these two antibody assays was found not to be optimal (Rho=0.79), as previously reported [18, 30-36], which is consistent with data showing that highly immunogenic epitopes within the S protein outside the RBD elicit potent NtAb responses [6,37].

The kinetics of SARS-CoV-2 RBD IgGs and NtAb followed a predictable course, as observed in previous publications [18,22,30-36], with antibody levels in both assays showing a consistent increase over time, and reaching a peak within the second and third week after the onset of symptoms for NtAb or slightly later for RBD-specific IgGs. Detection of NtAb at the early stages of COVID-19, irrespective of disease severity, has been previously reported [18,35]. By the end of the follow-up period more than two-thirds of patients in either ward had developed NtAb titers >1/160.

An interesting observation was that SARS-CoV-2 RBD IgGs avidity was quite low (<10%) in most sera, which were collected up to 2 months following the onset of symptoms, and showed minimal increase over time. This antibody avidity maturation pattern is reminiscent of that observed during SARS [38]. Remarkably, no correlation was found between SARS-CoV-2 RBD IgG AIs and NtAb_50_ titers. This finding is in agreement with the idea that limited to no affinity maturation is required from the germline to achieve a potent NtAb response to RBD [39].

The alleged association between high SARS-CoV-2 antibody levels and COVID-19 severity reported in a number of studies [17-22] is a matter of concern. If found to be the case, a plausible explanation for this observation may be that patients experiencing severe forms of the disease are exposed to higher and more perdurable viral burdens [18]; this, however, would call into question the role of antibodies in contributing to SARS-CoV-2 clearance. Alternatively, it may simply represent an epiphenomenom in the setting of an overall exaggerated immune response driven by “cytokine storms”, or may constitute a relevant pathogenetic mechanism involved in lung tissue damage (antibody-dependent enhancement) [15].

The data presented herein do not support the abovementioned association. In effect, we failed to find differences in SARS-CoV-2 RBD IgGs or SARS-CoV-2 NtAb_50_ levels within the first 30 days after the onset of symptoms between ICU and non-ICU patients who were matched for age, sex and co-morbidities. Furthermore, 2 out of the 4 ICU patients who died had relatively low NtAb_50_ titers (1/80). Liu and colleagues [19] showed that oxygen requirement in patients was independently associated with NtAb_50_ levels, as measured by both a pseudotyped reporter virus or live SARS-CoV-2 neutralization assay. Nevertheless, this finding should be interpreted with caution provided that only 8 ICU patients were recruited and these were much older than those in the non-ICU group. Wang et al. [18] also reported higher NtAb_50_ titers quantitated by a pseudotyped-virus based neutralization assay in severely ill patients as compared to mild COVID-19 patients. Interestingly, SARS-CoV-2 IgGs against S, S2, RBD and N were similar across groups. Unfortunately, no clinical characteristics of patients were reported other than the need for mechanical ventilation. Other studies including relatively small cohorts also pointed to an association of COVID-19 severity with SARS-CoV-2 NtAb [20,22,38]. In our view, comparison between studies addressing the abovementioned issue is rather problematic because of notable differences in clinical characteristics and therapeutic management of patients, categorization of severity, the timing of serum collection, and methods employed for SARS-CoV-2 antibodies detection and quantitation.

Disregulated synthesis and release of pro-inflammatory cytokines is thought to be a pathogenetic hallmark of most severe forms of COVID-19 [4-5]. Although the mechanisms of COVID-19–induced lung injury remain unclear, the so-called “cytokine storm” may likely play a critical role in the process of disease worsening and thus in COVID-19 prognosis [40]. Here, we investigated whether SARS-CoV-2 RBD IgG and NtAb_50_ levels correlate with serum concentrations of ferritin, Dimer-D, CRP, LDH and IL-6, which have been consistently shown to be markedly increased in patients with progressive disease and poor outcomes [4,5]. At most, we observed weak or very weak correlations between the antibody assays and these inflammatory biomarkers. Moreover, serum levels of the latter overlapped between patients with either high or low NtAb_50_ titers (≥ 1/160). Taken together, these data argue against a robust relationship between the magnitude of the antibody responses subjected to analysis herein and the state of inflammation in COVID-19 patients. To our knowledge, only one pre-print study used a similar approach to ours to address this issue [35], reporting a modest correlation (Rho=0.5) between NtAb_50_ titers and blood CRP levels. In addition, in contrast to what was observed here, a moderate negative correlation (Rho=-0.45) between NtAb_50_ titers and absolute lymphocyte counts was observed. As stated above, the comparison between the two studies is not straightforward.

The current study has several limitations. First, its retrospective nature. Second, cohort size is relatively small in our study. Third, IL-6 data was only available from 18 patients (all but one at ICU); in addition, all these patients were treated with tocilizumab. Fourth, SARS-CoV-2 antibodies and inflammatory biomarkers levels were measured in the blood compartment, which may not necessarily mirror those in lung tissue. Fifth, serum levels of other cytokines (i.e. TNF-α, or IL1-β) or chemokines (IFNγ-induced protein 10) that may reflect more accurately the overall state of inflammation were not measured [4,5]. Sixth, the data reported in the current study may be interpreted as arguing against a role for neutralizing antibodies in mediating SARS-CoV-2 clearance, as found in other studies that show an association between SARS-CoV-2 antibody levels and COVID-19. This would certainly be oversimplistic and against data published in experimental models [11]. Seventh, epitope specificities of SARS-CoV-2 antibodies other than for the S protein in the case of the neutralization assays or RBD in the case of the IgG tests were not assessed. In this sense, antibodies mediating immunopathogenetic events, especially through ADE, are more likely to behave as sub- or non-neutralizing and target epitopes outside RBD [4].

In summary, the data presented herein do not support an association between SARS-CoV-2 RBD IgG or NtAb_50_ levels and COVID-19 severity. Further, well-powered studies overcoming the abovementioned limitations are warranted to solve this question, which is of paramount relevance for vaccine design and for the safety of passive transfer therapies with plasma from convalescent COVID-19 individuals.

## Data Availability

The data that support the findings of this study are available on request from the corresponding author DN

## Acknowledgements

The members of the Decoy-SARS-CoV-2 Study Group from the Institute of Biomedicine of Valencia are the following ones: Vicente Rubio, Alberto Marina, Jeronimo Bravo, José Luis LLacer, Clara Marco, Alonso Felipe, Anmol Adhav, Carla Sanz, Nadine Gougeard, Susana Masiá, Francisca Gallego, Sara Zamora, Lidia Orea, Alicia Forcada, Alba Iglesias, Mónica Escamilla, Laura Villamayor, Borja Sáez, Carolina Espinosa and María Pilar Hernández. They form a group for production of proteins involved in SARS-COV-2 entry into cells and for analysis of their interactions. Their support as a team led by A. Marina was key to production of RBD protein used in the present study. The authors would like to thank Gert Zimmer (Institute of Virology and Immunology, Mittelhäusern/Switzerland), Stefan Pöhlmann and Markus Hoffmann (both German Primate Center, Infection Biology Unit, Goettingen/Germany) for providing the reagents required for the generation of VSV pseudotypes. Estela Giménez holds a Juan Rodés research contract from the Carlos III Health Institute (Ref. JR18/00053). Eliseo Albert holds a Río Hortega research contract from the Carlos III Health Institute (Ref. CM18/00221). Ron Geller holds a Ramón y Cajal fellowship from the Spanish Ministry of Economy and Competitiveness (RYC-2015-17517).

## Figure Legends

**Supplementary Figure 1.**
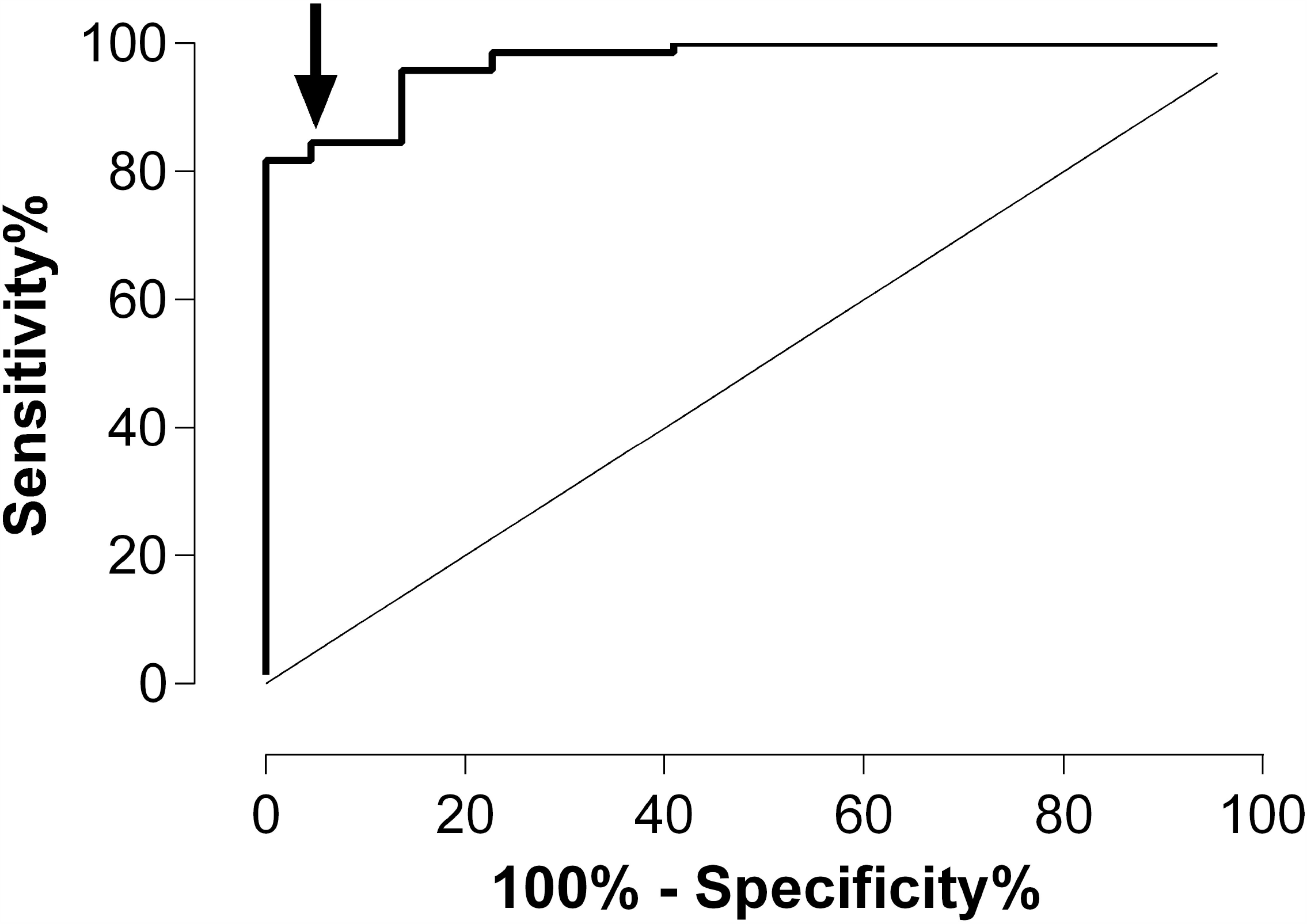
ROC curve analysis for establishing the optimal SARS-CoV-2 RBD IgG threshold level predicting the presence of high NtAb_50_ titers (≥ 1/160) in patients with COVID-19.

**Supplementary Figure 2.**
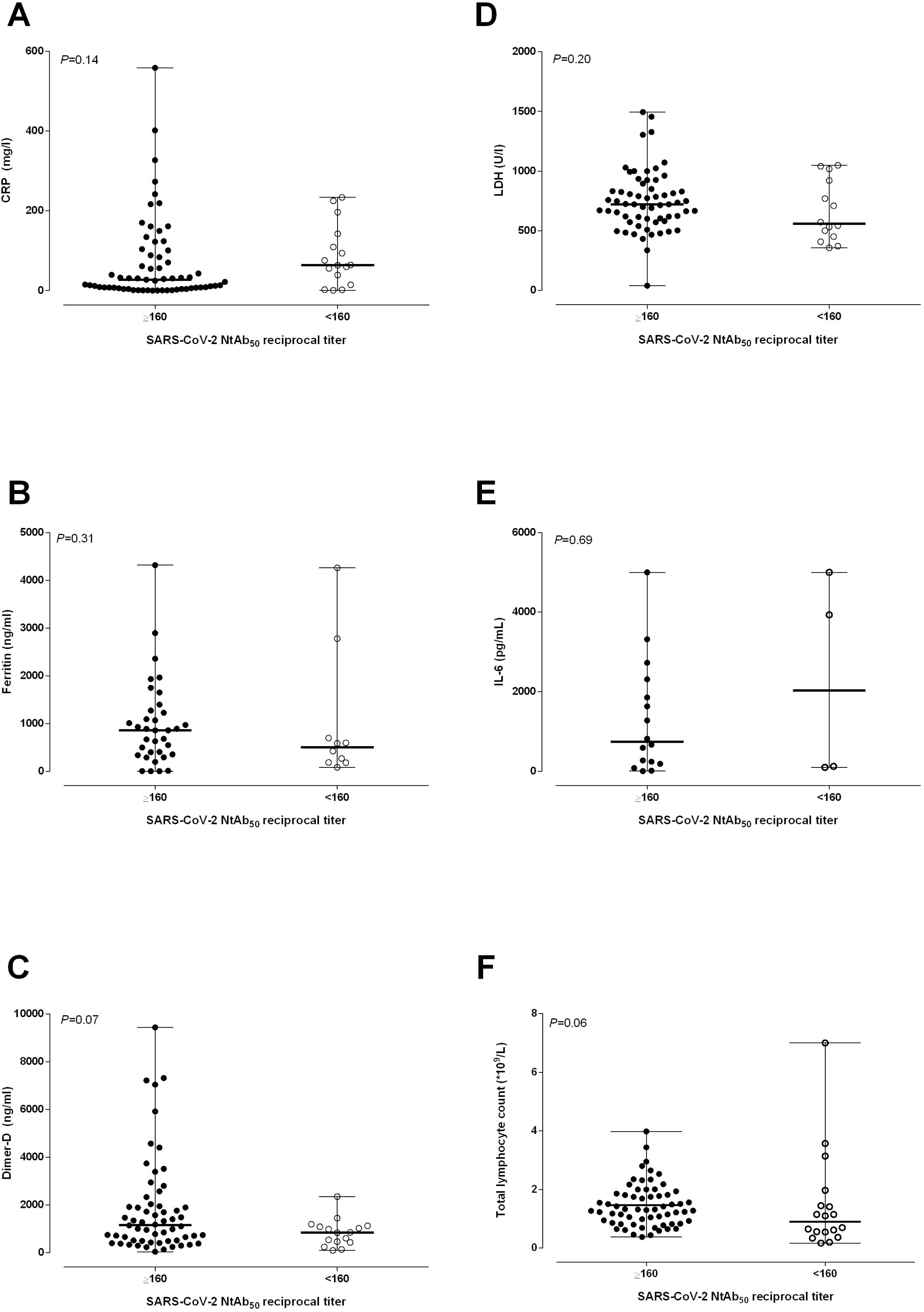
Serum levels of C-reactive protein (CRP), Dimer-D, ferritin, lactate dehydrogenase (LDH), interleukin-6 (IL-6) and absolute lymphocyte counts in COVID-19 patients with high (≥ 1/160) or low (<1/160) NtAb_50_ titers. *P* values for comparisons are shown.

